# Characterization of a fiber-coupled SPAD camera system for deep-tissue blood-flow measurement using diffuse correlation spectroscopy

**DOI:** 10.64898/2026.01.02.25343000

**Authors:** Christopher H. Moore, Michael A. Wayne, Arin C. Ulku, Paul Mos, Claudio Bruschini, Edoardo Charbon, Ulas Sunar

## Abstract

Diffuse correlation spectroscopy (DCS) is a promising technique for noninvasive measurement of blood flow, especially for cerebral blood flow where other noninvasive techniques have shortcomings. Conventional DCS often requires multiple simultaneous measurements to enhance the signal-to-noise ratio (SNR) especially when probing deep into the brain with large source-detector separations where photons are scarce. However, this limits scalability when using discrete optical detectors. This study demonstrates the application of the 500 x 500 single-photon avalanche diode (SPAD) array, SwissSPAD3, coupled with a custom field-programmable gate array (FPGA) design, which enables significant increases in SNR compared to conventional DCS systems. We validate the fiber-coupled SPAD camera system against a lab-standard CW-DCS system in two-layer liquid phantoms and in human measurements, and demonstrate robust blood-flow tracking at source-detector separations up to 3.25 cm. These results support SPAD-based parallel detection as a scalable route to improved deep-tissue DCS performance in humans.

## 1. Introduction

Diffuse correlation spectroscopy (DCS) is an optical, speckle-based technique for quantitative measurement of blood flow completely non-invasively. Traditional continuous-wave DCS (CW-DCS) involves the illumination of a diffusing sample, such as human tissue, with constant near-infrared coherent light like the output from a long-coherence laser [1–3]. The light diffuses through the medium and some of it returns to a single-mode fiber coupled to a detector that is able to measure the fluctuations of light on a scale of microseconds or faster. By measuring the temporal fluctuations in the speckle and performing an intensity autocorrelation to generate a g_2_ curve, we can determine its decorrelation time, which is inversely related to the movement speed of the scattering particles. The DCS technique has proven itself to be comparable to existing gold-standard, noninvasive quantitative measurement techniques including fMRI [4–8] and Doppler ultrasound [9]. With its growing popularity, DCS is finding applications in the assessment of cerebral activation [10–12], acute brain injuries [13–20], and peripheral arterial disease (PAD) [21,22]. However, with these applications comes the need to parallelize measurements to get multiple channels to obtain multiple source-detector separations and to generate maps of blood flow [23,24]. Additionally, some applications, especially cerebral ones due to the skull, require deeper probing in order to obtain a clinically useful signal. A simple way to do deeper probing is to increase the source-detector separation at the expense of losing much of the light returned to the detector which decreases signal-to-noise ratio (SNR). Alternatively, newer techniques such as time-domain DCS (TD-DCS) use a fast, pulsed laser to obtain a time point spread function (TPSF) where photons that travel deeper can be selected. However, this requires more complex fast, expensive detectors and more complex instrumentation overall [25–31].

Speckle contrast optical spectroscopy (SCOS) [32–35] operates on similar principles to DCS, but it does not rely on temporal speckle fluctuations observed by fast, expensive detectors. Instead, it uses standard CMOS or CCD camera sensors to capture speckle patterns produced by coherent light scattering from the sample. By tracking changes in the spatial distribution of these speckles over time, the decorrelation time can be calculated. The use of these cameras allows for a large detector surface area to be used with large-core fibers to deliver significantly more light from the sample to the detector compared to the single-mode fibers used for DCS. Since this technique allows for less expensive instrumentation, it is much more feasible to implement a multi-channel system by simply purchasing more cameras or dividing up a high-resolution sensor [34,36]. However, the standard SCOS technique is not a quantitative measurement of blood flow and most work done with SCOS will report normalized, relative changes. Compact SPAD-array implementations of SCOS and DCS have been demonstrated using low-density arrays (e.g. [37–39]).

Early multi-channel DCS systems were constrained by size and relied on multiple discrete detectors, each with its own fiber coupling to the sample, which quickly placed an upper limit on practical setups due to cost and space [40,41]. Cameras consisting of arrays of CMOS SPAD detectors have been developed more recently. This has allowed for improvements in SNR compared to single-detector systems by averaging multiple temporal speckle measurements [42–47]. These systems are able to achieve up to 500x increases in SNR over their single-pixel performance due to the massive number of parallel detectors and have pushed for larger source-detector separations as a result. Notably, Kreiss et al. used the 500×500 pixel SwissSPAD3 SPAD camera to achieve 4 cm source-detector separations while still maintaining enough temporal resolution to see pulsatility of the measured blood flow [42]. Additionally, the ATLAS SPAD presented a 512×512-pixel SPAD array with backside illumination that formed a 128×128-macropixel array where each macropixel had its own autocorrelator built into the CMOS chip to calculate the g_2_ in real time [45]. Recently, ATLAS-DCS has been used for deep non-invasive cerebral blood flow sensing, including pulsatile measurements at 50 mm source-detector separation in the human forehead [48]. In relation to these recent high-density SPAD-array DCS efforts, we focus here on a fiber-coupled, dual-separation SwissSPAD3 implementation with off-chip FPGA correlation. This enables simultaneous short- and long-separation measurements on a single sensor and direct benchmarking against a lab-standard CW-DCS system in phantoms and humans.

An issue faced when using DCS for deeper flow measurements is that the deep probing light also scatters through superficial tissue, thus contaminating the signal. This contamination means that the signal from the superficial tissue flow is also present in the signal of a long-separation probe that is trying to observe deeper flow. TD-DCS can overcome this as it filters out photons from superficial tissue, providing a measurement focused primarily on the deep tissue [27,31]. However, the complexity of the TD-DCS system and cost of the specialized individual detectors makes it a less effective solution. To overcome this issue for traditional CW-DCS, a solution being explored is to use a short-separation channel to measure the superficial signal at the same time as the long separation in order to use regression techniques to extract the flow dynamics that are exclusively in the deeper tissue [49,50].

The work presented here explores the application of the SwissSPAD3 SPAD camera for DCS in a multi-channel configuration [43,44]. Thanks to the large array of detector pixels provided by SwissSPAD3, the SNR of DCS measurements can be significantly improved to allow for increasingly large source-detector separations and make it possible to probe the blood flow deeper into the brain than conventional DCS systems. We show the use of a dual-channel system to enable simultaneous superficial and deep blood flow measurements with a single SPAD camera sensor. This system utilizes a highly sensitive long-separation channel for deep flow measurements alongside a less sensitive short separation to capture superficial flow, establishing a practical hardware approach to allow for potentially regressing out the confounding superficial signal. Our work emphasizes the quantitative experimental validation of this system against a lab-standard CW-DCS system, and showing robust in-vivo performance for human measurements.

## 2. Materials and Methods

### 2.1 Instrumentation

The setup used for our SPAD camera DCS measurements centers around the SwissSPAD3 binary-gated camera, a 500×500 array of SPAD detectors implemented in CMOS technology [44]. It has one of the highest pixel counts available for a SPAD array with a pixel pitch of 16.38 µm and active area of 6 µm diameter. We operated the camera at an operational voltage of 28V, corresponding to an excess bias voltage of 5V and a photon detection probability (PDP) of ∼13% at the 785nm wavelength we used for DCS. Sensor readout for each 250×500 pixel half of the sensor is performed by an OpalKelly XEM7360 FPGA development board hosting a Xilinx Kintex 410T FPGA. Due to the limited availability of these FPGA development boards, only the bottom 250×500 array of pixels could be used for these experiments.

The FPGA firmware reads in each 1-bit image from the camera sensor for the set exposure time and calculates 15 points of the g_2_ intensity autocorrelation by accumulating the recorded 1-bit images for the set integration time [43]. Fig. 1 shows the general workflow of this acquisition using the SPAD camera. The firmware is limited to a minimum exposure time of 10.81µs which also limits the minimum autocorrelation delay *τ*, calculated by the on-FPGA real-time correlators, to the same value, i.e. 10.81 µs. The range of autocorrelation delays can therefore be adjusted by changing the sensor exposure time and adding dummy exposures that allow the correlators to skip over unused *τ* values. However, for our work, this feature was not used, and the camera was set to always use an exposure of 10.81 µs and calculate 15 non-zero *τ* values linearly ranging from 10.81 µs to 162.15 µs to best capture fast flows with quickly decaying g_2_ curves. The integration time is also limited by the FPGA firmware to a maximum of 2^16^ exposures, corresponding to ∼0.71 s at 10.81 µs exposure time. This maximum value was used for all experiments reported below. The FPGA then sends the g_2_ curves over its USB3.0 interface to a computer for post-processing in MATLAB. Longer exposure times can be obtained by combining multiple integration times together in post-processing.

**Fig. 1.**
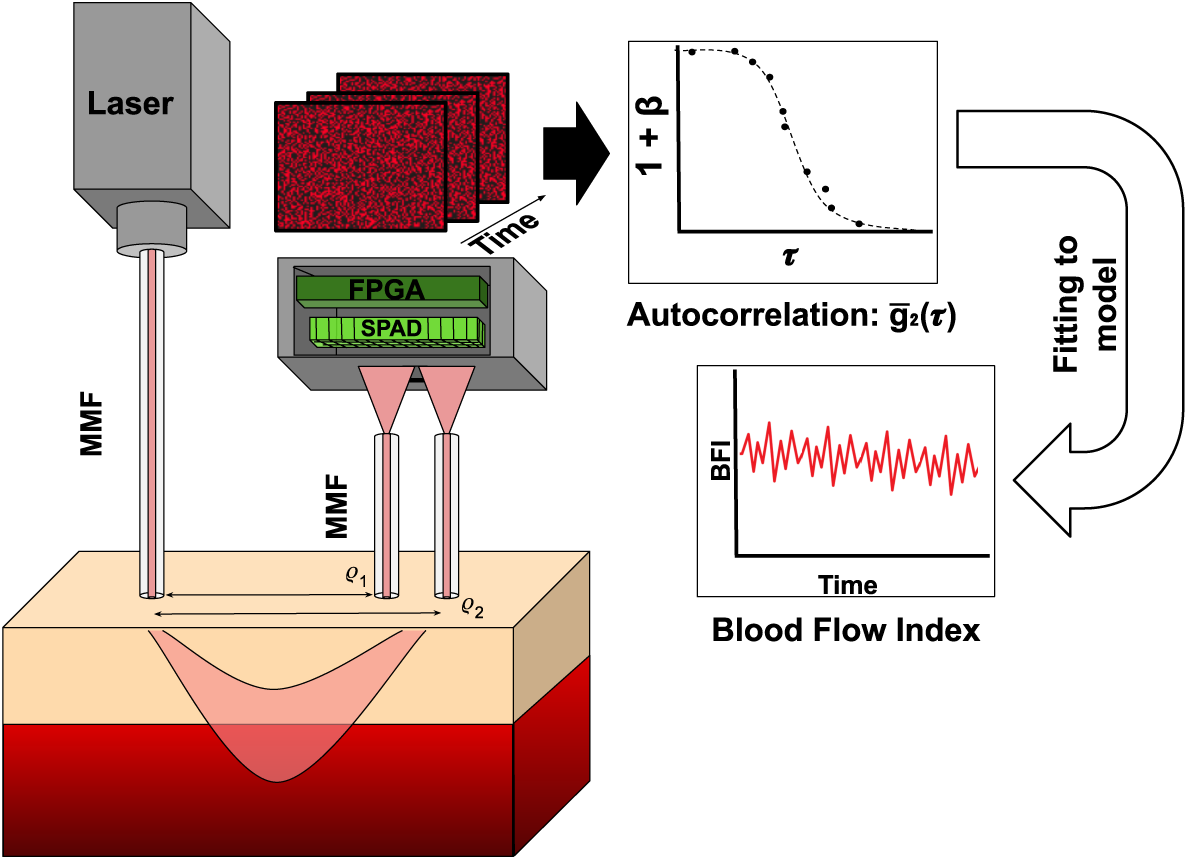
General workflow for performing DCS measurements with the SwissSPAD3 camera. A laser illuminates the sample through a multi-mode fiber, and multi-mode fibers collect the diffuse photons propagated through the medium. Each pixel of the SPAD acts as an individual detector for which the g2 is calculated. Pixels can then be either averaged together or individually fit to calculate a blood flow index (BFI) value.

Our setup butt-couples two 2 mm-diameter polymethyl methacrylate fibers (Super ESKA SH8001, Mitsubishi Chemical Corporation, Tokyo, Japan) to our SPAD array in order to obtain two separate channels while capturing as much light as possible from the illuminated sample. The two fibers are held by a custom 3D-printed mount that positions their tips in front of the bottom half of the sensor. A wall between the fibers extends to within a few millimeters of the sensor surface to prevent interference between the speckle patterns generated by each fiber, as shown in Figure 2. We adjusted the positioning of these two fibers such that one illuminated the majority of the half-sensor, while the other only illuminated a corner. The fiber illuminating the majority of the sensor is intended as a sensitive long-separation channel for deeper flow measurements without significant loss of SNR at lower photon fluxes. The other, less-sensitive channel is intended as a short-separation channel to allow for regression of the superficial signal from the long-separation channel. This short separation is subject to a higher photon flux and therefore does not need as many pixels to achieve high-SNR measurements. Due to the short separation’s higher photon flux, several film polarizers were stacked on top of each other at 90° angles to act as an adjustable filter so that the short separation would not saturate the sensor area that it illuminated, while still permitting it to operate at high count rates.

**Fig. 2.**
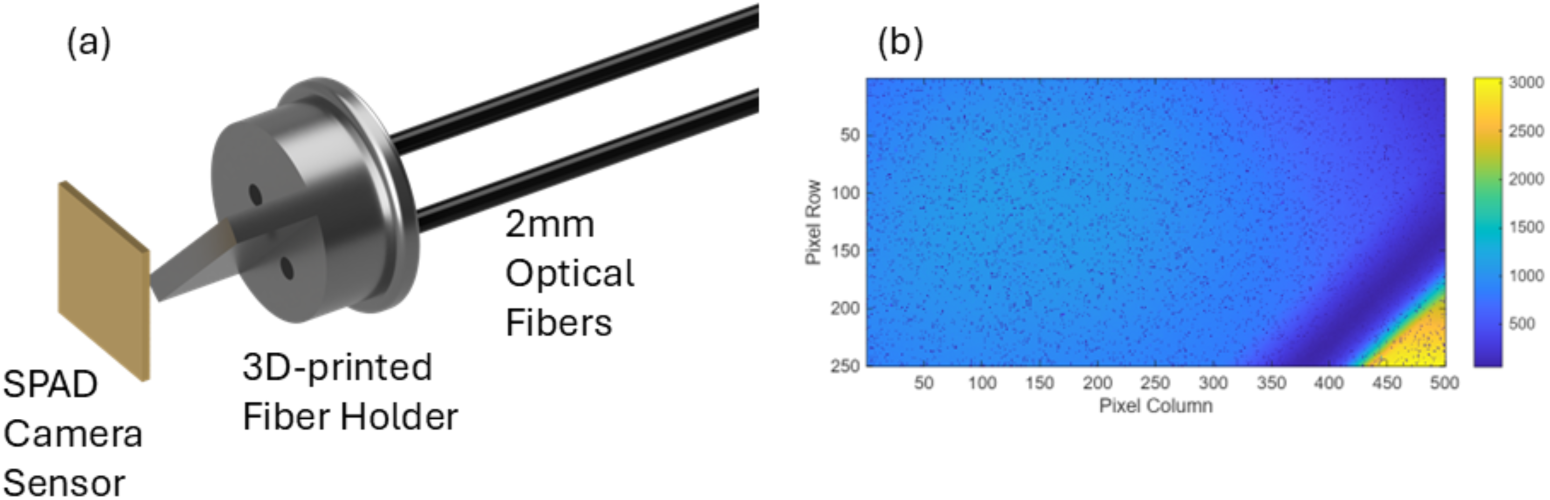
(a) A drawing of the 3D-printed fiber holder holding its two optical fibers in its approximate relative position to the SPAD camera’s sensor. (b) A sample image from the SPAD camera using the setup in (a) showing a large area of the half-sensor for the long separation, a shadow area from the divider wall, and a corner dedicated to the short separation.

The distances from the fiber tips to the sensor surface, z, were optimized according to Equation 1 where d is the average speckle diameter, *λ* is the wavelength of light, and D is the fiber core diameter [43]. The average speckle diameter was set to the same size as the pixel active area, so that each pixel measures approximately one speckle. Unlike SCOS which normally requires two pixels per speckle to use spatial sampling of the speckle and satisfy the Nyquist criterion [51], the DCS implemented with the SPAD camera uses temporal sampling of the speckle. Thus, the only requirement is that there are not multiple speckles in one pixel area. Using our fiber diameter of 2 mm, a of 785 nm, and speckle diameter of 6 μm, we calculated a fiber-tip-to-sensor distance of 15.3 mm. However, we ended up using a fiber-tip-to-sensor surface distance z of 20 mm to allow for the light exiting the fibers to spread and fully illuminate the sensor area. Although a setup using lenses to control the speckle size and area of the sensor covered by the fiber would be more flexible, we wanted to minimize losses from extra optical elements placed between the fiber tip and sensor surface.

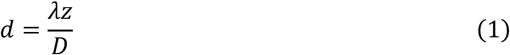

In order to average together and process the pixels for each of the two fiber channels separately, regions of interest (ROIs) were selected based on the intensity images generated by the camera. ROIs were selected by hand to ensure no overlap between the channels. This resulted in our larger, long-separation channel covering a region of 114,618 pixels or 91.7% of the half-sensor. The smaller, short-separation channel used a region of 7,975 pixels or 6.4% of the half-sensor. The remaining pixels, corresponding to the region shadowed by the wall in between the two fibers, were not used. Hot pixels were characterized by taking several dark images and placing a hot-pixel threshold at 10 median absolute deviations above the median intensity. This method labeled 4.1% of the total pixels for our half-sensor as hot ones, distributed evenly. For both channels, the g2 for the pixels from each sensor region excluding the dark pixels were averaged together into a single g2 curve for each channel at each time point.

The source for all experiments was a 785 nm continuous-wave laser (DL785-100, CrystaLaser Inc., Reno, NV, USA) coupled to a 600 µm multi-mode fiber. The power at the output of the multi-mode fiber was ∼70 mW. To validate our results and compare this SPAD camera system against lab-standard CW-DCS systems, we also used our lab’s established traditional CW DCS system using the same source laser with two avalanche photodiode detectors (SPCM-780-12-FC, Waltham, MA, USA) connected to a 4-channel hardware correlator (Flex01LQ-05, Correlator.com, Bridgewater, NJ, USA). Post-processing of both the SPAD camera and CW-DCS systems was done in MATLAB with both systems using identical code for the g_2_ fitting algorithm.

### 2.2 Two-Layer Phantom Experiments

Initial phantom experiments were done to objectively compare the outputs between the SPAD camera DCS and standard CW-DCS systems. To both evaluate the similarities between the two systems and the independence of the two channels on the SPAD camera, we made two-layer liquid phantoms. We 3D-printed a cylindrical container for the liquid phantom composed of two parts: a 75 mm-thick bottom cylinder to hold the bottom-layer liquid phantom, and an 11mm top ring to hold the top-layer liquid phantom and secure the fibers being used at different source-detector separations. A ∼40 µm-thick transparent plastic film was sandwiched between these two parts to prevent mixing of the top and bottom liquid phantom layers while still allowing light to pass between them.

We used two different liquid phantoms with this two-layer phantom container. A standard liquid phantom was made using diluted Intralipid (Fresenius Kabi, Uppsala, Sweden) and ink (Documental Eternal Ink, Higgins, Leeds, MA, USA) to create a phantom with an absorption coefficient (*µ_a_*) of 0.1 cm^−1^ and reduced scattering coefficient (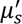) of 8 cm^−1^. This was designated as our “fast” phantom. We also made a “slow” phantom employing 30% glycerol (100% Pure Vegetable Glycerin, US+, New Haven, CT, USA). Using the adjustments in Intralipid concentration specified by Cortese et al. for glycerol phantoms [52], we set this slow phantom to the same *µ_a_* and 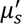 as our fast phantom. This 30% glycerol phantom is expected to have ∼45% less Brownian motion than the standard fast liquid phantom.

Using these two phantoms with our two-layer phantom container, we made a homogeneous two-layer phantom with both layers containing the fast liquid phantom (Fig. 3a) and a heterogeneous two-layer phantom with a slow top layer and fast bottom layer (Fig. 3b). The short separation was set to 0.8 cm and long separation to 3 cm for both the SPAD camera and CW-DCS systems. 300 integration times of 0.71 s were taken by each system and averaged together to generate a BFI for long and short separations in each phantom combination. We expected the homogeneous phantom to have minimal difference between the short and long separations. For the heterogeneous phantom, ideally, our short separation would see much slower flow due to the glycerol phantom, while the long separation remains unchanged compared to the homogeneous phantom. However, since the light for the long separation must first scatter through the slow top layer before reaching the fast bottom layer, both separations are expected to be slower. The long separation is still expected to be faster than the short one, but the difference between them may not amount to the full 45% difference as if the fast and slow phantoms were measured independently.

**Fig. 3.**
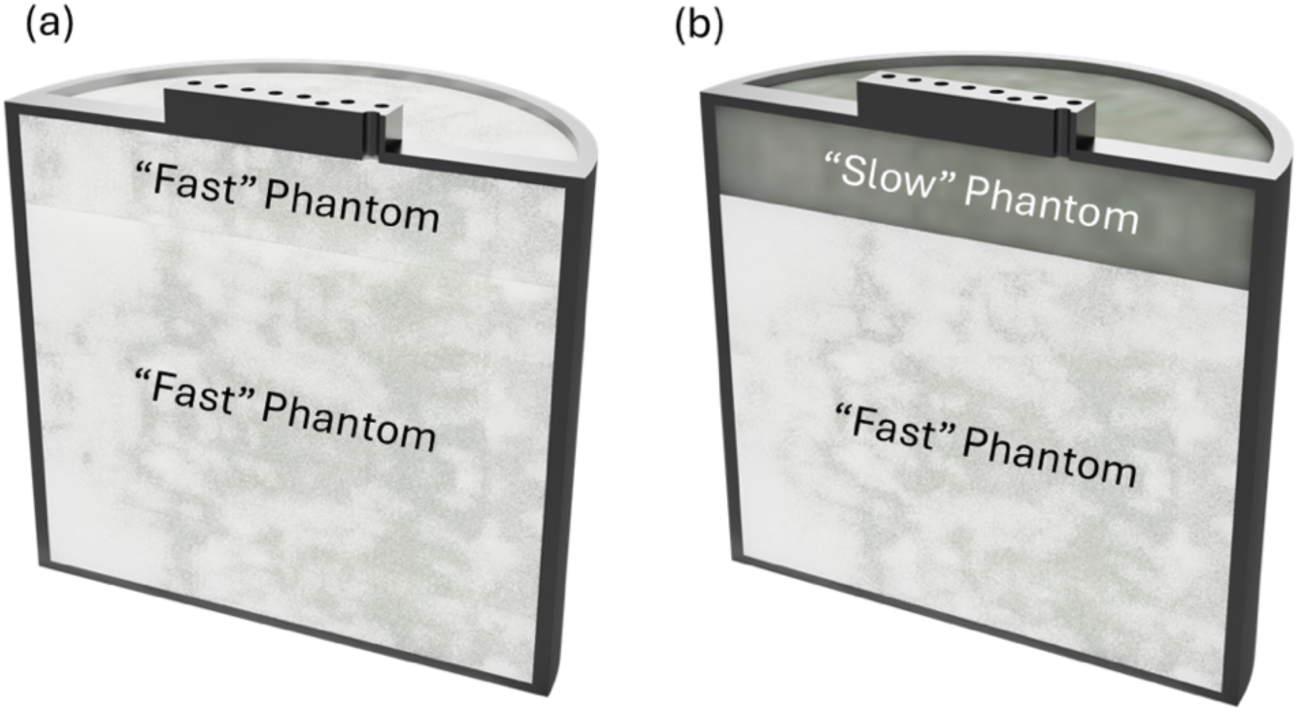
A cross section of the phantom container used for the two-layer phantom measurements. (a) The homogeneous phantom uses the same “fast” phantom to fill the top and bottom layers of the container. (b) The heterogeneous phantom keeps the “fast” phantom in the bottom layer, but replaces the top layer with the “slow” phantom.

### 2.3 SNR Characterization

We predicted that although each pixel detector has 57% lower detection efficiency (13% vs. 70%) and 27x worse timing precision (10.81µs minimum exposure vs. 400ns) compared to the single detectors of the CW system, the significantly improved light collection capabilities from using a SPAD camera with a large-core detector fiber (∼3 ∗ 10^$^ greater etendue over single-mode fiber) can have increasingly significant gains at large source-detector separations, where photons are scarce. To objectively evaluate the expected performance gains, we carried out SNR measurements using the same fast and slow liquid phantoms described before and calculated the resulting SNR using Equation 2 where *µ* and *σ* are the mean and standard deviation, respectively. The gain in SNR when using the camera’s long-separation channel was obtained by looking at the SNR of the lowest SwissSPAD3 *τ* of 10.81µs and comparing it to the CW-DCS system SNR using Equation 3. A value for the CW-DCS SNR at 10.81µs was generated by interpolating between the adjacent points since the CW-DCS hardware correlator does not generate a data point for the g_2_ at that time value.

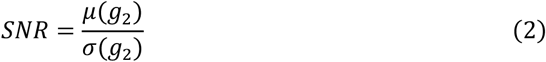

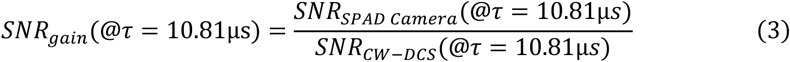

A standard phantom container was used to hold a single phantom at a time along with the detector fiber of the system under test. The laser source fiber was held in place by a linear motorized stage (MTS50-Z8) so that the source-detector separation could be adjusted by software. We used a MATLAB script to automatically take 60 integration times from the system being tested at 31 different source-detector separations, ranging from 2 cm to 4 cm with steps of 0.1 cm between 2 cm and 3 cm, then finer steps of 0.05 cm from 3 cm to 4 cm. This was repeated for the SPAD camera and CW-DCS systems with both the fast and slow phantoms. The average and standard deviation of the 60 integration times were calculated at each *τ* for the g2 curve to determine the SNR, though only the *τ* = 10.81*µs* point was used for the SNR gain calculation.

### 2.4 Human Measurements

All healthy participants provided written informed consent under a protocol approved by the Institutional Review Board (IRB) of Stony Brook University. Human measurements were performed using two standard techniques commonly used for evaluating DCS systems [25,28,53–59]. The first was an arm cuff ischemia experiment where we placed our 3D-printed DCS probe on the mid forearm and inflated a blood pressure cuff to 200mmHg to stop blood flow through the arm. We performed this experiment with a short 1cm separation and long 2.5cm separation. Our lab-standard CW-DCS system was used for simultaneous measurements as a comparison.

The intended application of this multi-channel SPAD camera DCS system is for deep blood flow measurement, one major physiological application being cerebral blood flow. To test the potential of our system for this application we performed breath hold experiments as our second standard technique. These experiments use the potent vasodilatory effects that increased CO_2_ in the blood has on cerebral vasculature. When the subjects hold their breath, their blood CO_2_ increases leading to a measurable increase in cerebral blood flow until the subject returns to normal breathing and the flow returns to baseline. We used a procedure identical to what was developed in Ozana et al. [59] to perform these experiments with a 1-minute baseline measurement, a 5 second breath out, hold for 25 seconds, and recovery for 2 minutes. Our DCS head probe was placed on the midline of the head on the forehead of the subject.

Since the work here pushes beyond the limits of traditional CW-DCS, no simultaneous CW-DCS was performed for the breath hold. We performed multiple trials of this breath hold experiment, each with a variable source-detector separation on the long-separation fiber to push its limits and a fixed 1cm separation on the short-separation fiber. We started at 3cm for the long separations, increasing the separation by 0.25cm for each trial until there was no longer any meaningful signal.

## 3. Results

### 3.1 Two-Layer Phantoms

Our first two-layer phantom was essentially homogenous with the fast liquid phantom being in both the top and bottom layers. Therefore, we would expect to see minimal changes between the long and short separations. We see this in the left side of Fig. 4 which shows these homogenous phantom results. The short and long separations for both systems are all within range of one standard deviation from each other. The right side of Fig. 4 shows the change we expected for the heterogeneous phantom with a slow top and fast bottom layer where the short separation shows much less flow than the long separations. These data confirm the validity of the measurements performed by our SPAD camera system against an established DCS standard. The result from the slow top with fast bottom liquid phantom also shows that we were able to successfully create two independent channels with our SPAD camera that are essentially indistinguishable from two separate detectors and do not have any noticeable crosstalk between them.

**Fig. 4.**
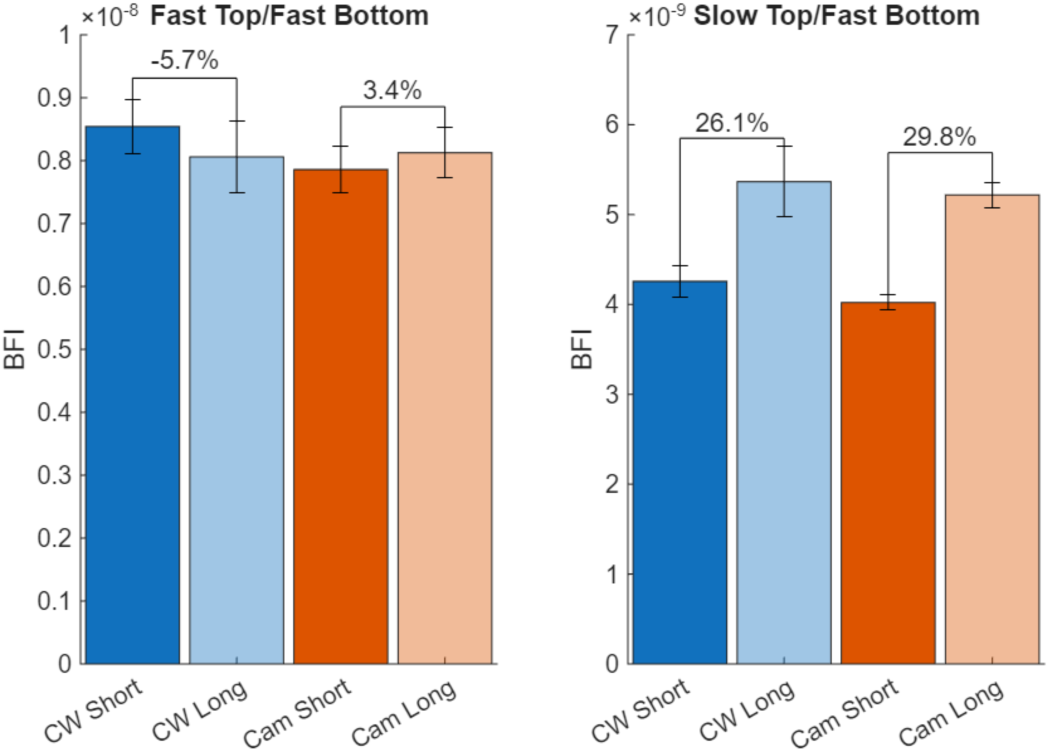
Bars represent the mean of the fit BFI for the short and long separations on the standard CW DCS and SPAD camera systems. Error bars show one standard deviation from the mean and the percent difference between the short and long separations are shown above the bars. The results from the homogeneous fast phantom are on the left and for the heterogeneous phantom with a slow top layer and fast bottom layer on the right.

### 3.2 Spatial Averaging Strategies

While the standard practice for processing multispeckle DCS g_2_ curves is by averaging them across the entire sensor or ROI prior to fitting, we also investigated whether subdividing the ROI and averaging the BFI post-fit could offer any improved robustness or error mitigation. The curve fitting procedure can introduce noise into the BFI estimation due to limitations in optimization accuracy, sensitivity of the cost function, and computational constraints even in the absence of strong noise in the g_2_ curves themselves [60,61]. By performing averaging after the fitting process, we could be more effectively using the many speckles we measure with the SPAD camera to average out the noise introduced from fitting. We evaluated the impact of different spatial averaging strategies by testing multiple grouping schemes using the already collected data from the long separation channel on our homogeneous liquid phantom data from the SPAD camera. We computed the standard deviation over time of BFI results obtained by first averaging across the full ROI prior to fitting and then dividing the ROI into increasing halved subgroups (2, 4, 8, …, 1024 groups) based on randomly selected pixels, fitting each group separately, and then averaging the results. Fig. 5 shows the results from this analysis and confirms the existing standard method of doing all averaging of the ROI prior to fitting. The standard deviation of the BFI increases as smaller ROI subdivisions are used, likely due to the noise of the g_2_ curve dominating over any noise introduced from the fitting process.

**Fig. 5.**
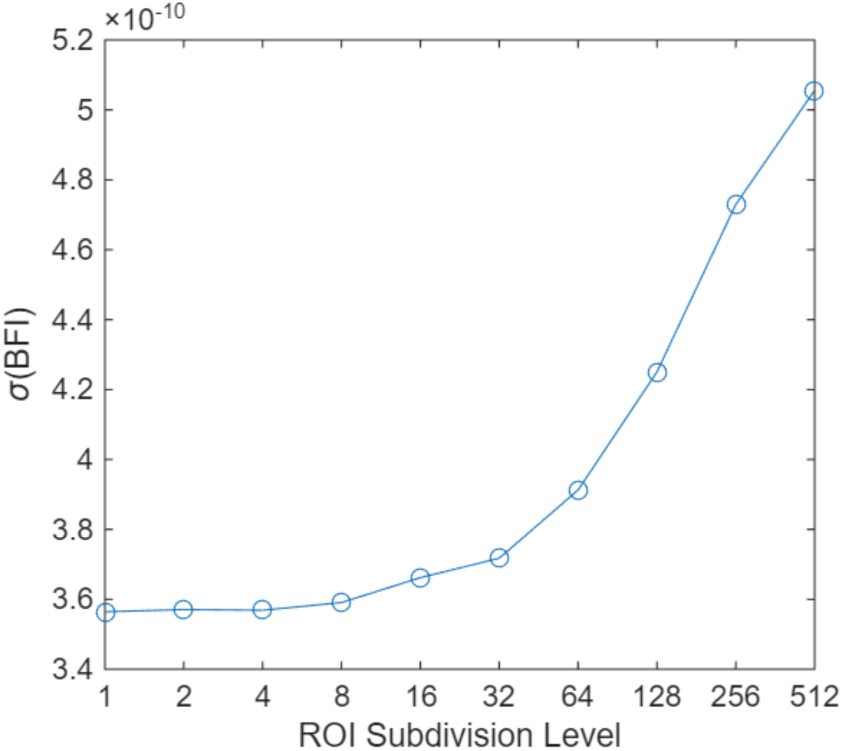
Standard deviation of fit BFI values as a function of the number of subdivisions of the ROI. The full ROI (subdivision level 1) was iteratively divided into random subset, doubling the number of groups at each step. For each configuration, g_2_ curves were averaged per group and then individually fit to extract BFI before averaging the groups together to obtain the final BFI for the selected time point. Standard deviation over the experiment time period was then calculated from these final BFI values.

### 3.3 SNR Characterization

Data from the characterization of the fast liquid phantom are shown here first with Fig. 6 which is SPAD camera SNR data. We see that the source-detector separation of 2.7 cm shows the highest overall SNR with SNR dropping off going either closer or further away. This can be attributed to the saturation of the SPAD camera at separations lower than 2.7 cm. Even though there is higher photon flux and we expect there to be higher SNR, the camera is unable to read and reset the pixels fast enough, so photons pile up and reduce SNR. We also see the expected drop in SNR as *τ* increases since SNR decreases as the g_2_ decays.

**Fig. 6.**
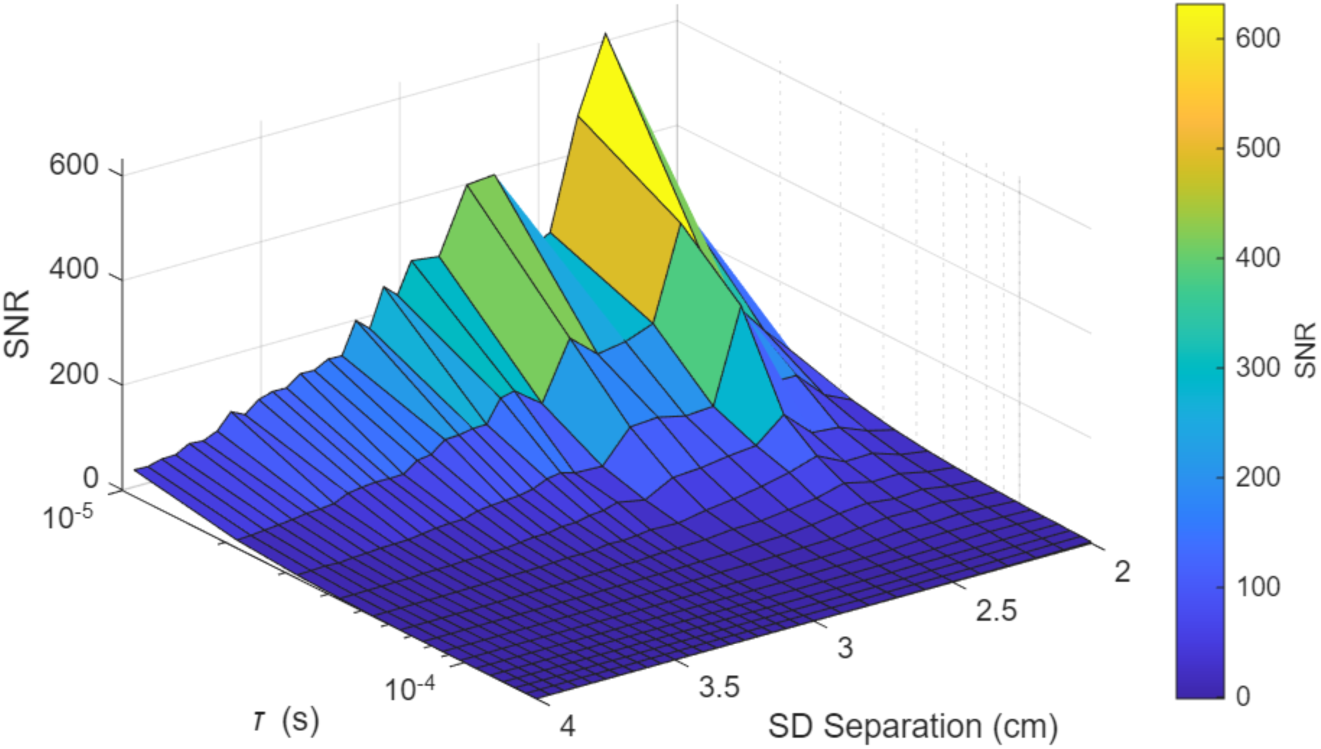
Surface plot of the SPAD camera SNR for fast liquid phantom at source-detector separations ranging from 2 cm to 4 cm across the full range of *τ* that the camera can generate.

Fig. 7 shows the equivalent data from the CW-DCS system. Since this system is able to capture *τ* values down to 400ns and over a larger range up to 1s, we see a more prevalent peak near the minimum *τ* and at the minimum separation measured of 2cm instead of a larger separation. The single detector from this system is able to sustain much higher count rates so it does not saturate as easily as the SPAD camera and since it has a faster response, the minimum *τ* is more than an order of magnitude smaller than for the SPAD camera. The smaller minimum *τ* allows us to see where the g_2_ flattens out at the top, where the signal is at its maximum. Since we do not see this peak in the SNR of the SPAD camera plot, it is reasonable to say that increasing the frame rate (e.g., by reading out a subset of the rows) of the camera would allow for the SNR of the SPAD camera to be further maximized.

**Fig. 7.**
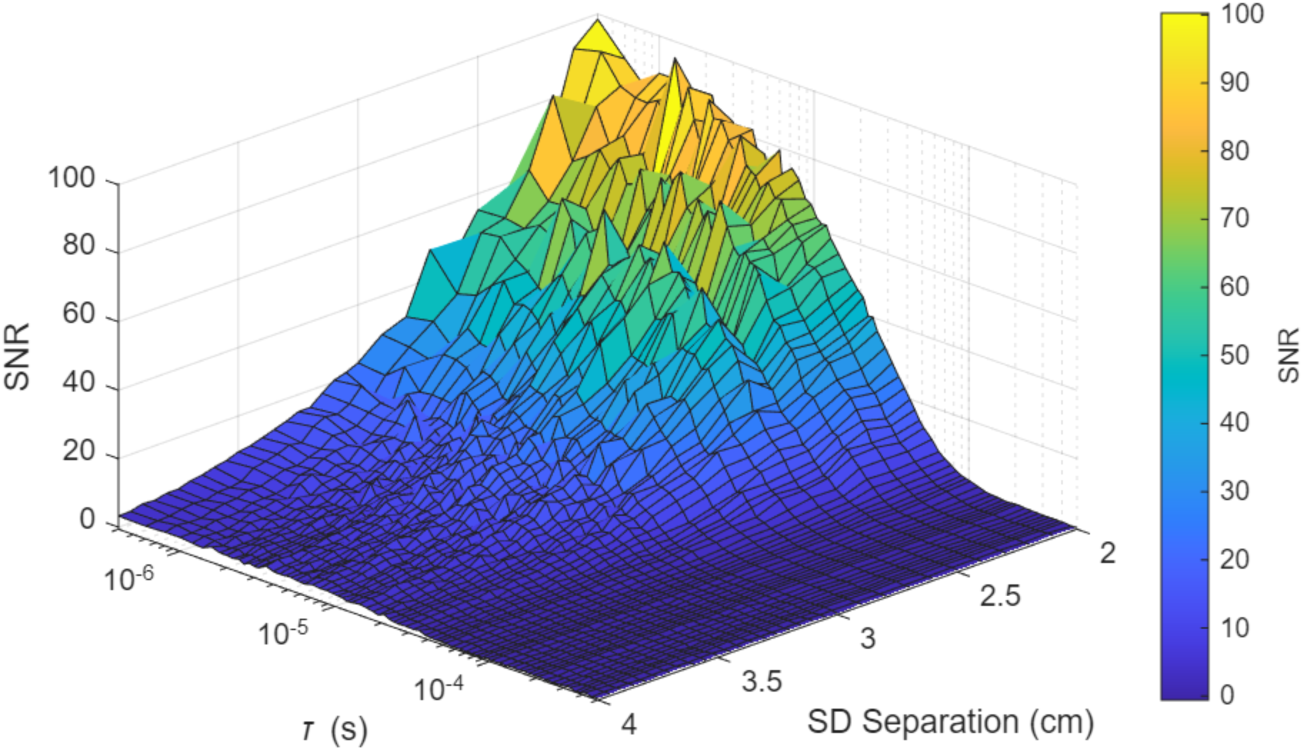
Surface plot of the CW DCS SNR for fast liquid phantom at source-detector separations ranging from 2 cm to 4c m. Although the CW DCS system can generate *τ* up to nearly one second, this plot only shows up to 5*10^-4^ seconds as SNR is essentially constant past this region.

There is a clear increase in SNR when using the SPAD camera long separation compared to the CW system, as we would indeed expect from the significant increase in the number of detectors provided by the SPAD camera (Fig. 8). However, we also see that the SNR gain further increases as source-detector separation increases. This was initially unexpected as the multispeckle DCS noise model would predict a constant gain in SNR across the source-detector separation range, assuming that the photon count rate decreased proportionally for both systems [46]. Further investigation into this showed that the photon count rate decreased for the CW DCS system faster than for the SPAD camera. This demonstrates the ability of our camera system to better capture light in photon-starved environments.

**Fig. 8.**
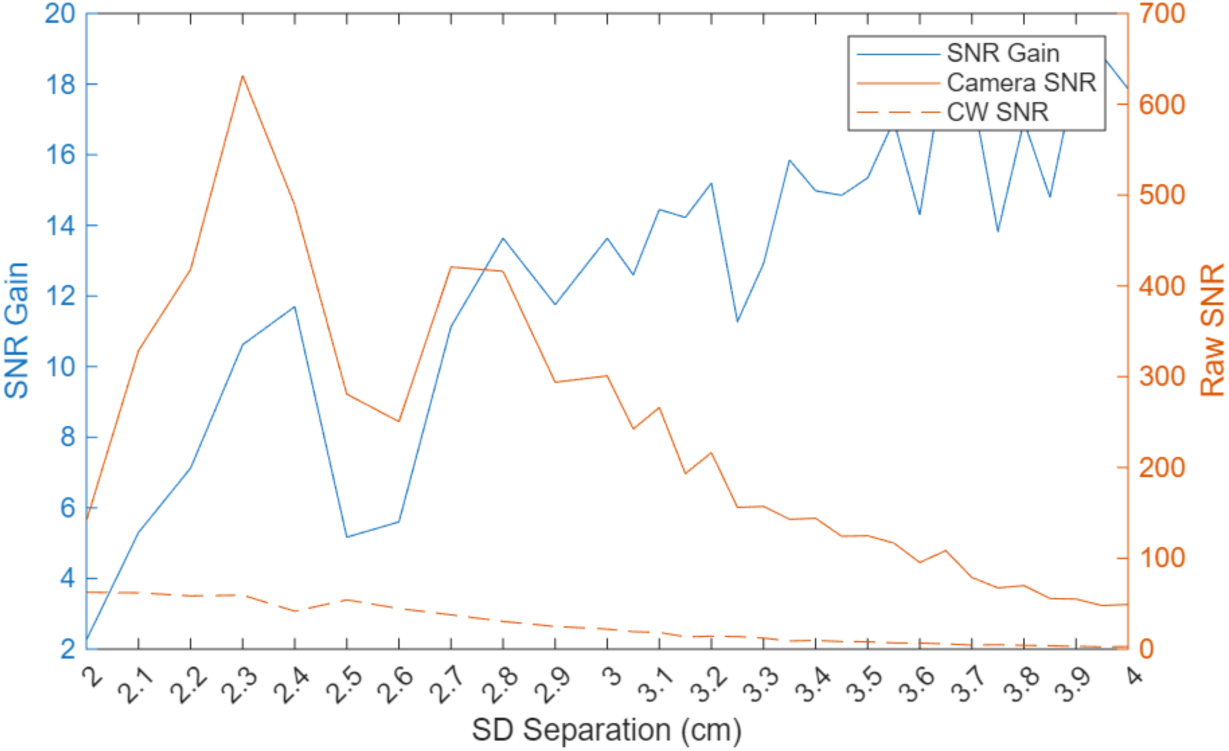
A direct comparison of the SNRs for the SPAD camera and CW DCS systems at *τ* = 10.81*µs* for the fast liquid phantom. The SNR gain is the ratio of the camera SNR to the CW SNR.

Results from the slower glycerol liquid phantom measurements are largely similar to fast liquid phantom. Although the slower phantom should have allowed for the limited range of *τ* from the SPAD camera, we are still not able to capture an obvious peak in the SPAD camera SNR shown in Fig. 9 like we can for the CW-DCS system in Fig. 10. Fig. 9 continues to demonstrate the saturation of the SPAD camera at separations below 2.7 cm as the SNR drops with anything closer.

**Fig. 9.**
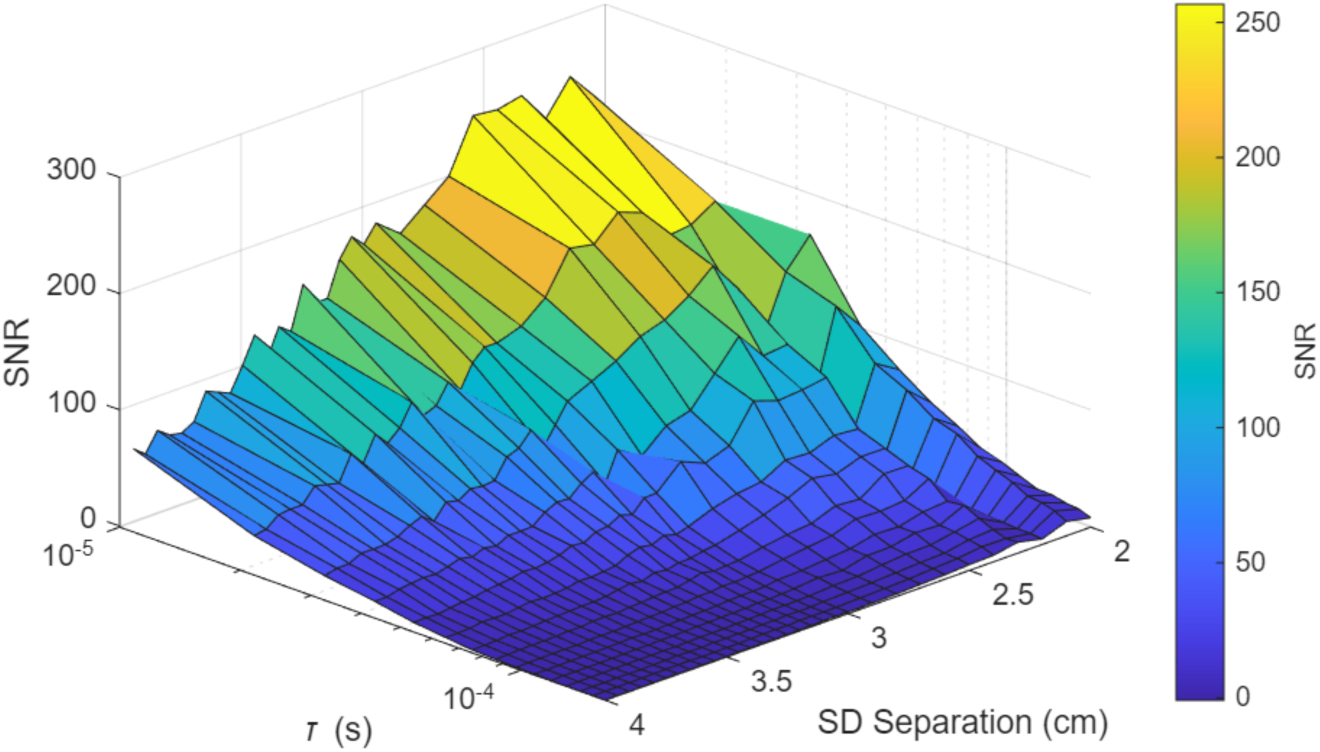
Surface plot of the SPAD camera SNR for slow glycerol liquid phantom at source-detector separations ranging from 2 cm to 4cm across the full range of *τ* that the camera can generate.

**Fig. 10.**
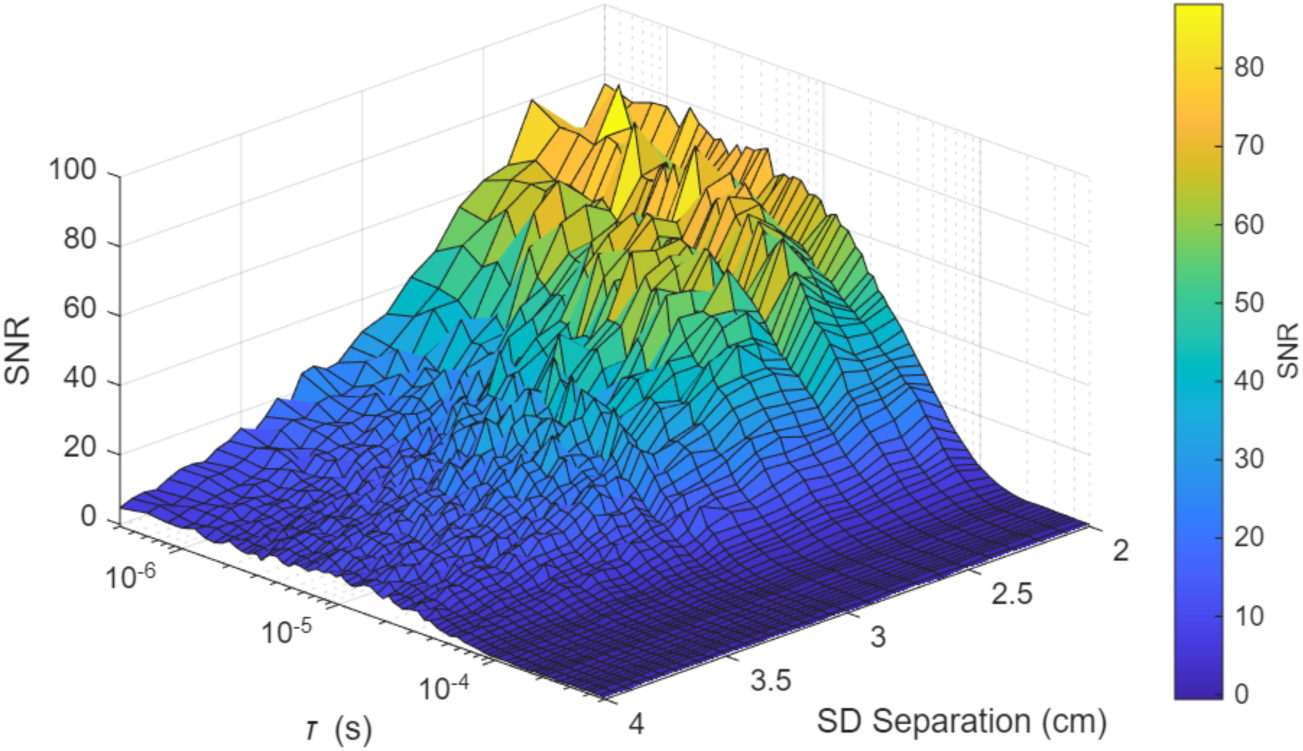
Surface plot of the CW DCS SNR for slow glycerol liquid phantom at source-detector separations ranging from 2 cm to 4cm. Although the CW DCS system can generate *τ* up to nearly one second, this plot only shows up to 5*10^-4^ seconds as SNR is essentially constant past this region.

Fig. 11 continues to show consistent results with the SNR gain given by the camera increasing as source-detector separations increase and photons get scarce.

**Fig. 11.**
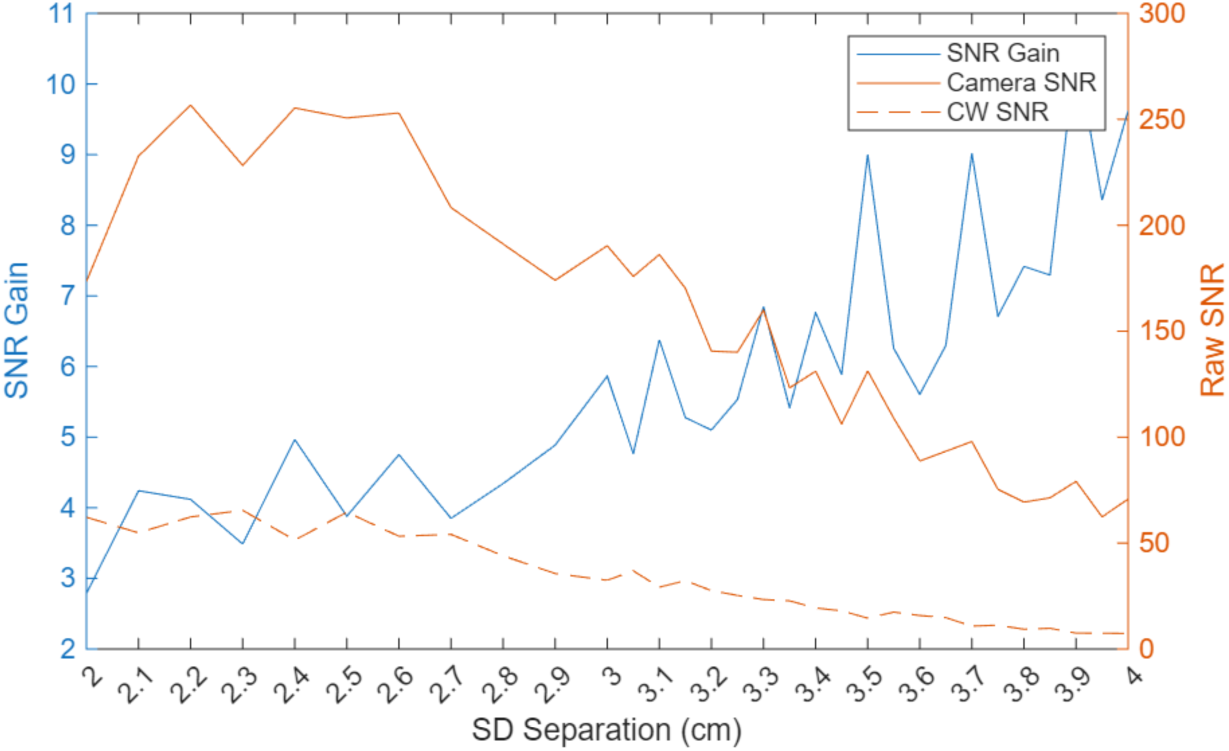
A direct comparison of the SNRs for the SPAD camera and CW DCS systems at *τ* = 10.81*µs* for the slow glycerol liquid phantom. The SNR gain is the ratio of the camera SNR to the CW SNR.

### 3.4 Human Measurements

Raw BFI results from the cuff ischemia experiment comparing the camera and CW-DCS systems are shown in Fig. 12. The initial baseline period from 0s to 60s shows a strong match between the respective channels on each system. There is a transient rise in BFI coincident with cuff inflation immediately before the onset of occlusion. This feature is most likely dominated by inflation associated mechanical perturbations, including minor probe motion, although a physiological contribution from discomfort related muscle activation cannot be fully excluded. During the occlusion from 60s to 120s we see the expected sharp drop in BFI followed by a reactive hyperemia as the cuff is released, with the short separations showing more dramatic changes in BFI. During the recovery period, the short separations do not fully return to baseline while the long separations do. This behavior is consistent with the greater sensitivity of short separations to superficial tissue hemodynamics and to local probe pressure and coupling effects. Most importantly, the SPAD camera measurements closely track the temporal dynamics shown by the established CW system and capture the contrast between the long and short separations.

**Fig. 12.**
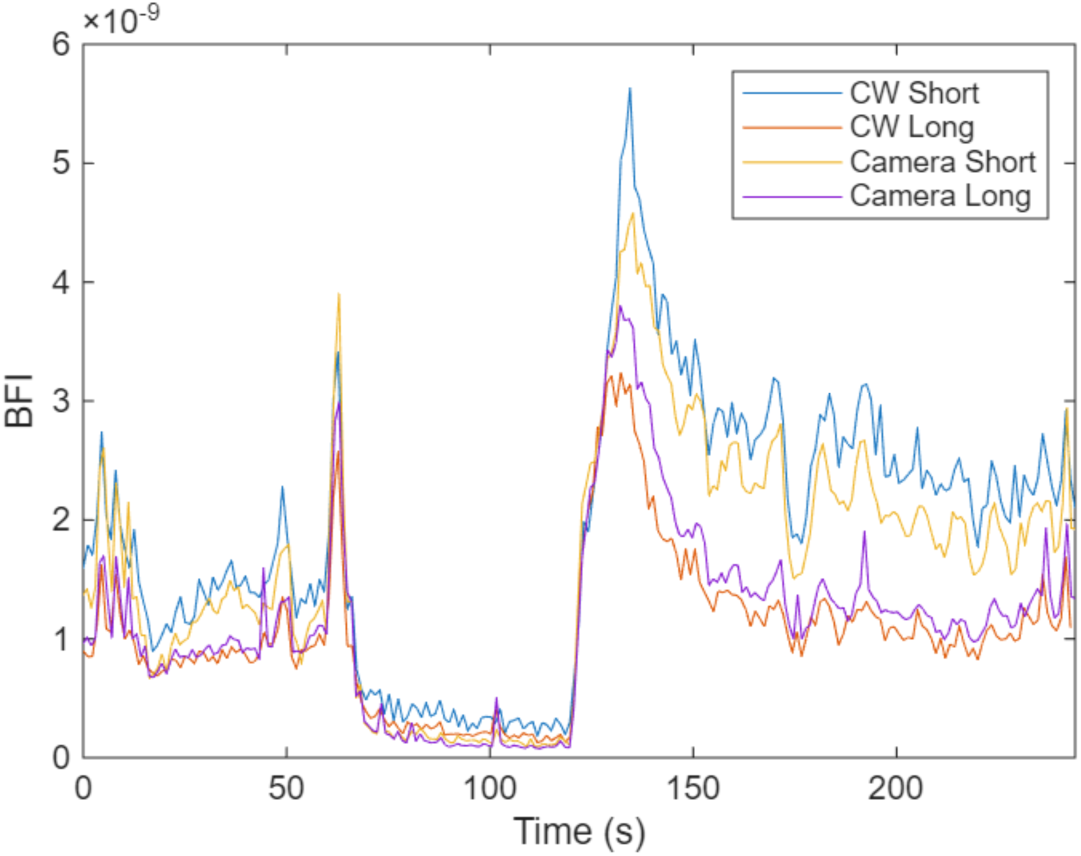
Comparison of BFI measured during a cuff ischemia protocol using the SPAD camera system and our lab’s established CW DCS system. Measurements are done with a short 1 cm and long 2.5 cm separations on both systems. We see the expected drop in BFI followed by a peak as the cuff is released. The SPAD camera closely matches the results from the CW system in both separations.

To more objectively compare the results from these two systems, we performed a Bland-Altman analysis on each channel as shown in Fig. 13. For the short separation the mean bias was small relative to the actual BFI being measured and had a symmetric distribution around the mean, without any obvious trends in bias as the BFI increases. This suggests a good agreement between the systems at the short separation. The long separation showed a smaller mean bias, although there is a more obvious trend of increasing discrepancy with higher BFI. This could be attributed to increasing error introduced from the fitting process as the BFI gets higher due to the SPAD camera having so few points calculated in its g_2_ curve and the minimum *τ* being high compared to the CW system. Despite these biases, most of the data points for both separations are within the 1.96 standard deviation limits, indicating that the two systems return comparable results across this range.

**Fig. 13.**
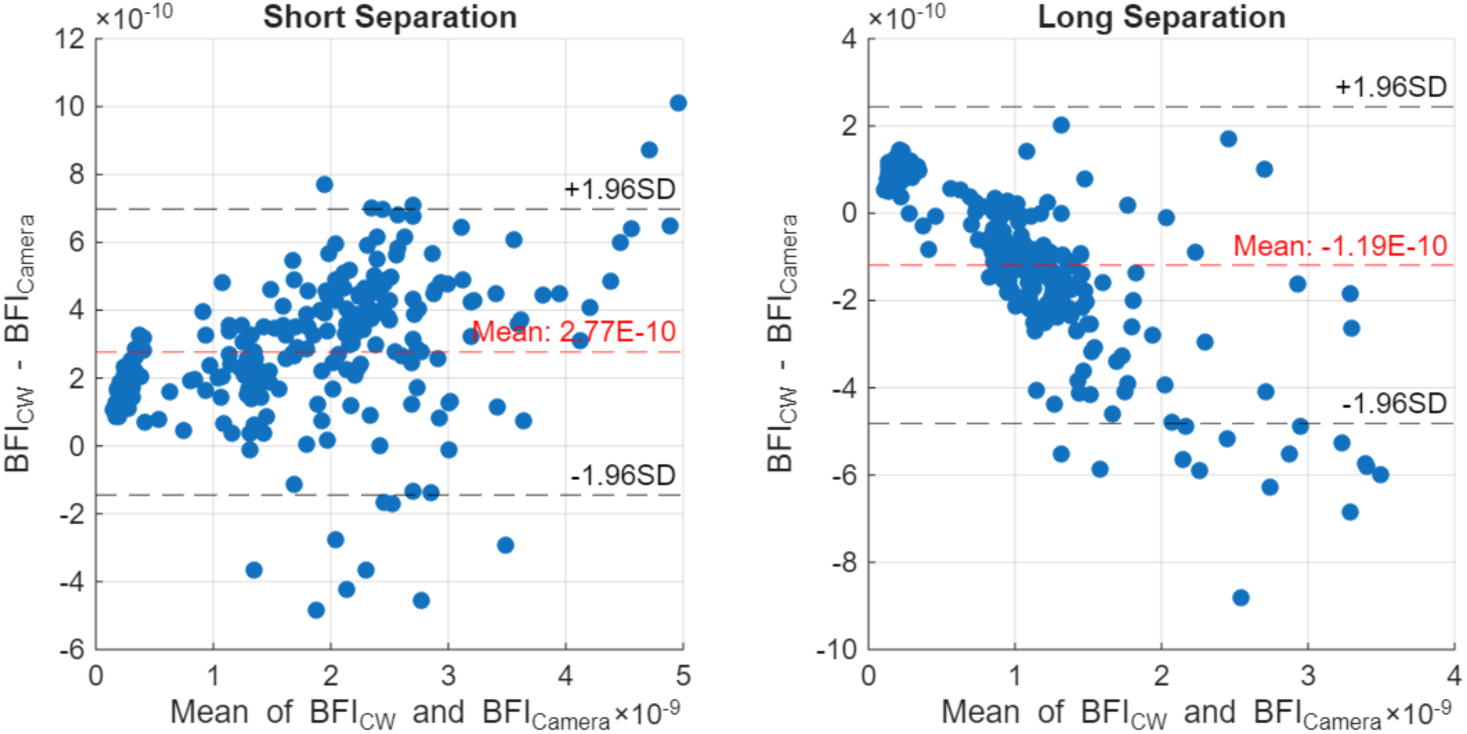
Bland-Altman plots comparing SPAD camera and CW BFI measurements for short (left) and long (right) separations. The red dashed line indicates the mean difference (bias), and black dashed lines show the ±1.96 standard deviation (SD) limits. At short separation, the bias is minimal with an even distribution of points, suggesting strong agreement between the systems. At long separation, a slight negative bias and increasing spread suggest that deeper measurements show greater variability.

The goal with our breath-hold experiments was to push the limits on source-detector separation with our camera system. Simultaneous CW system measurements were thus not performed as it was unable to return any meaningful data at such large separations. We were able to achieve a separation of 3.25 cm while still getting g_2_ data which we could fit without any temporal averaging or filtering. The BFI results from this 3.25 cm experiment are shown in Fig. 14. Both long and short separations show a gradual increase in BFI during the breath hold at around the 60 s timepoint. This is followed by a return to baseline after normal breathing resumes. Previous literature shows that the short separation should show a slightly larger and delayed change compared to the long separation, which is not the case here [59]. However, the previously shown breath-hold BFI changes were not significantly different over much of the breath hold time period and the observed changes were highly variable based on the subject, which may explain these differences. Despite this, our work’s focus is to push for deeper measurements with high SNR which we successfully show here.

**Fig. 14.**
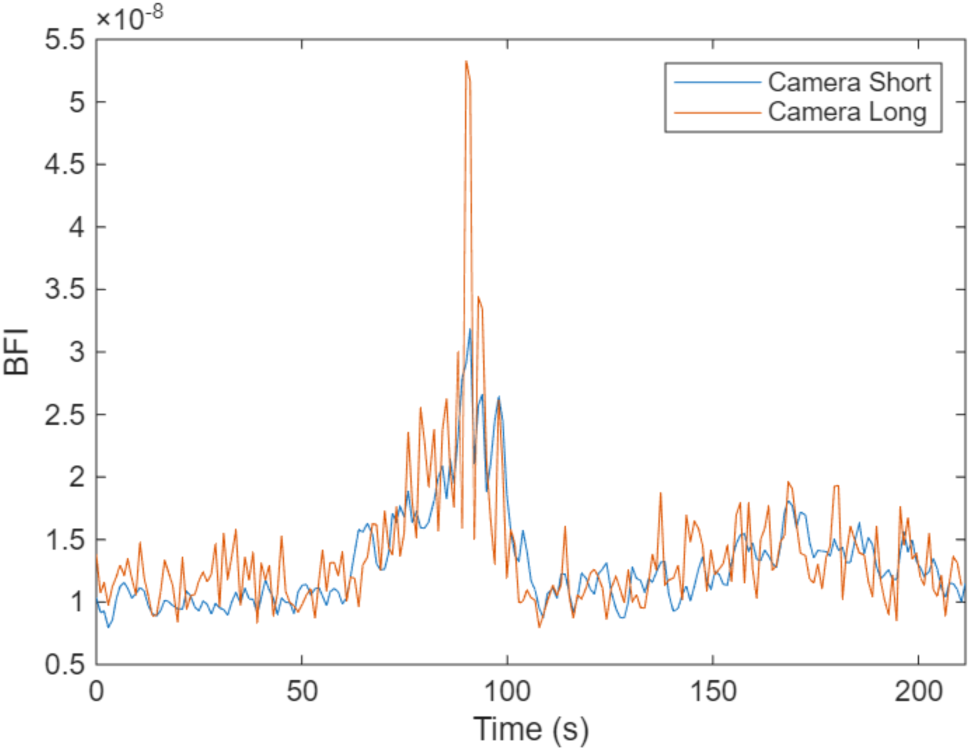
BFI responses measured by the SPAD camera system during a breath hold experiment with a short separation of 1 cm and long separation of 3.25 cm. Both short and long source-detector separations show an increase in BFI during the breath hold period, with the long separation signal showing a more pronounced peak, although generally matching the short separation.

## 4. Discussion

We have successfully demonstrated in this work the implementation of a dual-channel, multi-separation DCS system using the SwissSPAD3 CMOS SPAD camera. With the large array of detector pixels, we were able to achieve significantly enhanced performance for deep blood flow imaging compared to conventional DCS systems. By leveraging our ability to use large core detector fibers and parallelization, we were able to maintain strong SNR at large source-detector separations while the CW-DCS system started to struggle due to a lack of photons.

Relative to previous SPAD camera-based DCS implementations, our work demonstrates a validated dual-channel architecture which enables taking independent short and long-separation measurements and explicitly targeting the potential to separate superficial and deep flow contributions to the long-separation signal. Ultimately, this dual-channel framework provides a practical path toward isolating deep flow contributions, which may yield more physiologically relevant measurements than approaches that rely solely on extending source-detector separation, where superficial flow contamination remains a fundamental limitation. Furthermore, our approach performs autocorrelation on an external FPGA and leverages flexible ROI selection for multi-fiber channelization on a single sensor.

Our two-layer phantom experiments confirmed our systems’ ability to independently resolve superficial and deep flow across separate channels. The homogeneous phantom first confirmed agreement between the long and short separations across both systems, while the heterogeneous phantom demonstrated the sensitivity of the long separation to deep layers. Importantly, our SPAD camera system yielded results that match well with our lab-standard CW-DCS system. Our human measurements further reinforced this agreement between systems with the cuff-ischemia results showing similar temporal dynamics between the SPAD and CW systems at both separations. Bland-Altman analysis confirmed generally tight agreement for short separations and some bias for long separations. The slight divergence at high BFI in long-separation data may stem from the SPAD camera’s limited autocorrelation point density and relatively high minimum *τ*, which becomes more significant during fast flow conditions where decorrelation occurs more rapidly.

One of the clear advantages of the SPAD camera system is shown by the SNR characterization across varying source-detector separations. The ability of the SPAD camera to average over many speckles showed increasingly larger SNR gain over the CW system, especially beyond 2.7 cm separations. This contrasts with the DCS noise model, which predicts constant SNR gain assuming proportional photon loss across systems. Instead, we found that the SPAD camera suffered less photon loss than the CW-DCS system with increasing separation, highlighting its superior light collection efficiency via large-core fibers and wide-area detection.

Our most clinically relevant result was our ability to perform breath-hold experiments at a source-detector separation of 3.25 cm, which is a separation at which our conventional CW DCS system was unable to return meaningful data. At this separation, the SPAD camera was able to maintain sufficient SNR to track cerebral blood flow dynamics without the need for temporal averaging or post-processing filters. This reinforces the SPAD camera’s potential as a platform for deep brain flow monitoring in clinical applications, especially where standard CW systems are photon-limited. While the short and long-separation channels during breath hold showed similar trends, future work can be done to better isolate the deep BFI signal using short-separation regression techniques. The two-channel setup shown here provides the groundwork for that approach, and our success in decoupling phantom layers already indicates that these channels are sufficiently independent for regression analysis.

There are several limitations in the current implementation of the SPAD camera. First, the SPAD camera is limited in the range of the g_2_ curve that it is able to calculate. The minimum *τ* was limited to 10.81 µs primarily by the camera’s frame rate and the FPGA-based correlator is limited to 15 autocorrelation points. These limits primarily make it more difficult for the SPAD camera to accurately resolve fast flows, but very slow flow can also be affected. Although its current state is sufficient for most physiological measurements, improvements in the camera’s timing are required to show greater ranges of BFI. They would likely also benefit the accuracy of measurements even in the current range of BFIs. The sensitivity of the SPAD pixels at the core of SwissSPAD3 are also limiting for human applications, due to the detection efficiency at the NIR wavelengths typically used for DCS. The photon detection efficiency at the 785nm wavelength is indeed low compared to typical single-detector DCS system, but as more work shows the effectiveness of using 1064 nm sources for cerebral blood flow measurements [31], pushing for higher wavelengths will only make this detection efficiency worse for silicon-based platforms. A SPAD camera designed specifically for NIR wavelength applications could further highlight the benefits of using SPAD cameras. Autocorrelation bias in SPAD-array DCS at fast sampling times and in low-photon regimes has been characterized, along with correction strategies that may further improve quantitative accuracy [62].

The two-channel design demonstrated here has the potential to be expanded further into many more channels to generate tomographic images. The SPAD array already has an ample number of detectors to perform long separation measurements and adding another FPGA to our SwissSPAD3 would allow us to unlock the other, currently unused, half of the SPAD camera sensor. A multi-fiber, multi-separation probe could be made using a single SPAD camera to enable tomographic flow imaging and the use of regression techniques could further enhance the separation of flow at different depths in the resulting flow images.

Our work establishes the feasibility of a dual-channel, multi-separation DCS system using the SwissSPAD3 SPAD camera to substantially enhance deep blood flow imaging. The demonstrated capability to maintain high SNR at larger source-detector separations, coupled with robust agreement between our SPAD camera and established CW-DCS systems in both phantom and human studies, illustrates the system’s potential for clinical applications, particularly in deep brain monitoring. Although limitations such as the restricted range of the g₂ curve and sensitivity challenges at NIR wavelengths currently constrain the system’s performance, these findings provide a solid foundation for future improvements. By expanding the two-channel design into a multi-channel and eventually a tomographic framework, and by addressing technical constraints, we envision a new generation of DCS systems that can deliver detailed and depth-resolved blood flow measurements in photon-limited environments.

## Data availability

The data in this study are not publicly available but may be obtained from the corresponding author upon reasonable request.

## Acknowledgements

The authors acknowledge the funding support from NIH R01 (NIBIB Brain Initiative (7R01EB031759-03). ChatGPT (GPT-5) was used to assist with grammar and language editing during the preparation of this manuscript.

## References

1. R. C. Mesquita, T. Durduran, G. Yu, E. M. Buckley, M. N. Kim, C. Zhou, R. Choe, U. Sunar, and A. G. Yodh, “Direct measurement of tissue blood flow and metabolism with diffuse optics,” Philosophical Transactions of the Royal Society A: Mathematical, Physical and Engineering Sciences 369, 4390–4406 (2011).

2. T. Durduran and A. G. Yodh, “Diffuse correlation spectroscopy for non-invasive, micro-vascular cerebral blood flow measurement,” Neuroimage 85, 51–63 (2014).

3. T. Durduran, R. Choe, W. B. Baker, and A. G. Yodh, “Diffuse optics for tissue monitoring and tomography,” Reports on Progress in Physics 73, 076701 (2010).

4. A. G. Yodh, T. F. Floyd, C. Zhou, G. Yu, J. Wang, J. A. Detre, and T. Durduran, “Validation of diffuse correlation spectroscopy for muscle blood flow with concurrent arterial spin labeled perfusion MRI,” Optics Express, Vol. 15, Issue 3, pp. 1064–1075 15, 1064–1075 (2007).

5. T. Durduran, C. Zhou, E. M. Buckley, M. N. Kim, G. Yu, R. Choe, J. William, G. Thomas, L. Spray, S. M. Durning, S. E. Mason, L. M. Montenegro, S. C. Nicolson, R. A. Zimmerman, M. E. Putt, J. Wang, J. H. Greenberg, J. A. Detre, A. G. Yodh, and D. J. Licht, “Optical measurement of cerebral hemodynamics and oxygen metabolism in neonates with congenital heart defects,” 10.1117/1.3425884 15, 037004 (2010).

6. G. P. Dai, Y. R. Kim, M. A. Franceschini, S. A. Carp, and D. A. Boas, “Validation of diffuse correlation spectroscopy measurements of rodent cerebral blood flow with simultaneous arterial spin labeling MRI; towards MRI-optical continuous cerebral metabolic monitoring,” Biomedical Optics Express, Vol. 1, Issue 2, pp. 553–565 1, 553–565 (2010).

7. G. P. Dai, Y. R. Kim, M. A. Franceschini, S. A. Carp, and D. A. Boas, “Validation of diffuse correlation spectroscopy measurements of rodent cerebral blood flow with simultaneous arterial spin labeling MRI; towards MRI-optical continuous cerebral metabolic monitoring,” Biomedical Optics Express, Vol. 1, Issue 2, pp. 553–565 1, 553–565 (2010).

8. G. Yu, T. F. Floyd, T. Durduran, C. Zhou, J. Wang, J. A. Detre, and A. G. Yodh, “Validation of diffuse correlation spectroscopy for muscle blood flow with concurrent arterial spin labeled perfusion MRI,” Opt Express 15, 1064 (2007).

9. C.-G. Bangalore-Yogananda, R. Rosenberry, S. Soni, H. Liu, M. D. Nelson, and F. Tian, “Concurrent measurement of skeletal muscle blood flow during exercise with diffuse correlation spectroscopy and Doppler ultrasound,” Biomed Opt Express 9, 131 (2017).

10. J. Li, T. Elbert, J. Kissler, M. Ninck, L. Koban, and T. Gisler, “Transient functional blood flow change in the human brain measured noninvasively by diffusing-wave spectroscopy,” Optics Letters, Vol. 33, Issue 19, pp. 2233–2235 33, 2233–2235 (2008).

11. J. Li, F. Jaillon, T. Elbert, T. Gisler, and G. Dietsche, “Activity of the human visual cortex measured non-invasively by diffusing-wave spectroscopy,” Optics Express, Vol. 15, Issue 11, pp. 6643–6650 15, 6643–6650 (2007).

12. A. G. Yodh, C. Zhou, J. H. Greenberg, G. Yu, J. A. Detre, J. Wang, T. Durduran, and M. G. Burnett, “Diffuse optical measurement of blood flow, blood oxygenation, and metabolism in a human brain during sensorimotor cortex activation,” Optics Letters, Vol. 29, Issue 15, pp. 1766–1768 29, 1766–1768 (2004).

13. C. S. Poon, B. Rinehart, D. S. Langri, T. M. Rambo, A. J. Miller, B. Foreman, and U. Sunar, “Noninvasive Optical Monitoring of Cerebral Blood Flow and EEG Spectral Responses after Severe Traumatic Brain Injury: A Case Report,” Brain Sciences 2021, Vol. 11, Page 1093 11, 1093 (2021).

14. M. T. Mullen, A. B. Parthasarathy, A. Zandieh, W. B. Baker, R. C. Mesquita, C. Loomis, J. Torres, W. Guo, C. G. Favilla, S. R. Messé, A. G. Yodh, J. A. Detre, and S. E. Kasner, “Cerebral Blood Flow Response During Bolus Normal Saline Infusion After Ischemic Stroke,” Journal of Stroke and Cerebrovascular Diseases 28, 104294 (2019).

15. M. N. Kim, B. L. Edlow, T. Durduran, S. Frangos, R. C. Mesquita, J. M. Levine, J. H. Greenberg, A. G. Yodh, and J. A. Detre, “Continuous optical monitoring of cerebral hemodynamics during head-of-bed manipulation in brain-injured adults,” Neurocrit Care 20, 443–453 (2014).

16. D. R. Busch, R. Balu, W. B. Baker, W. Guo, L. He, M. Diop, D. Milej, V. Kavuri, O. Amendolia, K. St. Lawrence, A. G. Yodh, and W. A. Kofke, “Detection of Brain Hypoxia Based on Noninvasive Optical Monitoring of Cerebral Blood Flow with Diffuse Correlation Spectroscopy,” Neurocrit Care 30, 72–80 (2019).

17. W. B. Baker, R. Balu, L. He, V. C. Kavuri, D. R. Busch, O. Amendolia, F. Quattrone, S. Frangos, E. Maloney-Wilensky, K. Abramson, E. Mahanna Gabrielli, A. G. Yodh, and W. Andrew Kofke, “Continuous non-invasive optical monitoring of cerebral blood flow and oxidative metabolism after acute brain injury,” Journal of Cerebral Blood Flow and Metabolism 39, 1469–1485 (2019).

18. R. M. Forti, M. Katsurayama, J. Menko, L. Valler, A. Quiroga, A. L. E. Falcão, L. M. Li, and R. C. Mesquita, “Real-Time Non-invasive Assessment of Cerebral Hemodynamics With Diffuse Optical Spectroscopies in a Neuro Intensive Care Unit: An Observational Case Study,” Front Med (Lausanne) 7, 503559 (2020).

19. R. M. Forti, C. G. Favilla, J. M. Cochran, W. B. Baker, J. A. Detre, S. E. Kasner, M. T. Mullen, S. R. Messé, W. A. Kofke, R. Balu, D. Kung, B. A. Pukenas, N. I. Sedora-Roman, R. W. Hurst, O. A. Choudhri, R. C. Mesquita, and A. G. Yodh, “Transcranial Optical Monitoring of Cerebral Hemodynamics in Acute Stroke Patients during Mechanical Thrombectomy,” Journal of Stroke and Cerebrovascular Diseases 28, 1483–1494 (2019).

20. J. Selb, K.-C. Wu, J. Sutin, P.-Y. Lin, P. Farzam, S. Bechek, A. Shenoy, A. B. Patel, D. A. Boas, M. A. Franceschini, and E. S. Rosenthal, “Prolonged monitoring of cerebral blood flow and autoregulation with diffuse correlation spectroscopy in neurocritical care patients,” Neurophotonics 5, (2018).

21. R. C. Mesquita, M. P. Malavika, C. Guoqiang, Y. Xiaoman, X. Sung, W. Han, G. Lech, Y. Shang, T. Durduran, C. Zhou, A. G. Yodh, E. R. Mohler, M. Putt, M. Chandra, G. Yu, X. Xing, S. W. Han, and E. R. M. Iii, “Diffuse optical characterization of an exercising patient group with peripheral artery disease,” 10.1117/1.JBO.18.5.057007 18, 057007 (2013).

22. G. Yu, T. Durduran, G. Lech, C. Zhou, B. Chance, E. R. M. M.D., and A. G. Yodh, “Time-dependent blood flow and oxygenation in human skeletal muscles measured with noninvasive near-infrared diffuse optical spectroscopies,” 10.1117/1.1884603 10, 024027 (2005).

23. R. Paul, K. Murali, and H. M. Varma, “High-density diffuse correlation tomography with enhanced depth localization and minimal surface artefacts,” Biomed Opt Express 13, 6081 (2022).

24. S. Han, J. Johansson, M. Mireles, A. R. Proctor, M. D. Hoffman, J. B. Vella, D. S. W. Benoit, T. Durduran, and R. Choe, “Non-contact scanning diffuse correlation tomography system for three-dimensional blood flow imaging in a murine bone graft model,” Biomed Opt Express 6, 2695 (2015).

25. S. Wojtkiewicz, M. Kacprzak, N. Mogharari, D. Borycki, and A. Liebert, “Time-domain diffuse correlation spectroscopy at large source detector separation for cerebral blood flow recovery,” Biomedical Optics Express, Vol. 15, Issue 7, pp. 4330–4344 15, 4330–4344 (2024).

26. V. Parfentyeva, L. Colombo, P. Lanka, M. Pagliazzi, A. Brodu, N. Noordzij, M. Kolarczik, A. Dalla Mora, R. Re, D. Contini, A. Torricelli, T. Durduran, and A. Pifferi, “Fast time-domain diffuse correlation spectroscopy with superconducting nanowire single-photon detector: system validation and in vivo results,” Sci Rep 13, 1–11 (2023).

27. J. Sutin, B. Zimmerman, D. Tyulmankov, D. Tamborini, K. C. Wu, J. Selb, A. Gulinatti, I. Rech, A. Tosi, D. A. Boas, and M. A. Franceschini, “Time-domain diffuse correlation spectroscopy,” Optica 3, 1006 (2016).

28. N. Ozana, N. Lue, M. Renna, M. B. Robinson, A. Martin, A. I. Zavriyev, B. Carr, D. Mazumder, M. H. Blackwell, M. A. Franceschini, and S. A. Carp, “Functional Time Domain Diffuse Correlation Spectroscopy,” Front Neurosci 16, 932119 (2022).

29. S. Samaei, P. Sawosz, M. Kacprzak, Ż. Pastuszak, D. Borycki, and A. Liebert, “Time-domain diffuse correlation spectroscopy (TD-DCS) for noninvasive, depth-dependent blood flow quantification in human tissue in vivo,” Sci Rep 11, 1–10 (2021).

30. J. Sutin, I. Rech, B. Zimmerman, D. Tyulmankov, M. A. Franceschini, J. Selb, D. Tamborini, K. C. Wu, D. A. Boas, A. Tosi, and A. Gulinatti, “Time-domain diffuse correlation spectroscopy,” Optica, Vol. 3, Issue 9, pp. 1006–1013 3, 1006–1013 (2016).

31. C.-S. Poon, D. S. Langri, B. Rinehart, T. M. Rambo, A. J. Miller, B. Foreman, and A. U. Sunar, “First-in-clinical application of a time-gated diffuse correlation spectroscopy system at 1064&#x2005;nm using superconducting nanowire single photon detectors in a neuro intensive care unit,” Biomedical Optics Express, Vol. 13, Issue 3, pp. 1344–1356 13, 1344–1356 (2022).

32. “Comparing the performance potential of speckle contrast optical spectroscopy and diffuse correlation spectroscopy for cerebral blood flow monitoring using Monte Carlo simulations in realistic head geometries,” https://www.spiedigitallibrary.org/journals/neurophotonics/volume-11/issue-01/015004/Comparing-the-performance-potential-of-speckle-contrast-optical-spectroscopy-and/10.1117/1.NPh.11.1.015004.full.

33. C.-H. P. Lin, C.-H. P. Lin, I. Orukari, C. Tracy, L. K. Frisk, M. Verma, S. Chetia, T. Durduran, T. Durduran, J. W. Trobaugh, J. P. Culver, and J. P. Culver, “Multi-mode fiber-based speckle contrast optical spectroscopy: analysis of speckle statistics,” Optics Letters, Vol. 48, Issue 6, pp. 1427–1430 48, 1427–1430 (2023).

34. B. Kim, S. Zilpelwar, E. J. Sie, F. Marsili, B. Zimmermann, D. A. Boas, and X. Cheng, “Measuring human cerebral blood flow and brain function with fiber-based speckle contrast optical spectroscopy system,” Commun Biol 6, 1–10 (2023).

35. R. Bi, K. Lee, and J. Dong, “Deep tissue flowmetry based on diffuse speckle contrast analysis,” Optics Letters, Vol. 38, Issue 9, pp. 1401–1403 38, 1401–1403 (2013).

36. C. Luk, C. Luk, C.-H. P. Lin, C.-H. P. Lin, F. Beslija, M. Verma, L. K. Frisk, S. Chetia, T. Durduran, T. Durduran, J. P. Culver, J. P. Culver, E. Richter, and J. W. Trobaugh, “Multi-Mode Fiber-Based Speckle Contrast Optical Tomography System for Humans,” Optica Biophotonics Congress 2025 (2025), paper JM4A.7 JM4A.7 (2025).

37. T. Dragojević, J. L. Hollmann, D. Tamborini, D. Portaluppi, M. Buttafava, J. P. Culver, F. Villa, and T. Durduran, “Compact, multi-exposure speckle contrast optical spectroscopy (SCOS) device for measuring deep tissue blood flow,” Biomed Opt Express 9, (2018).

38. J. D. Johansson, D. Portaluppi, M. Buttafava, and F. Villa, “A multipixel diffuse correlation spectroscopy system based on a single photon avalanche diode array,” J Biophotonics 12, (2019).

39. T. Dragojević, D. Bronzi, H. M. Varma, C. P. Valdes, C. Castellvi, F. Villa, A. Tosi, C. Justicia, F. Zappa, and T. Durduran, “High-speed multi-exposure laser speckle contrast imaging with a single-photon counting camera,” Biomed Opt Express 6, 2865 (2015).

40. C. J. Stapels, N. J. Kolodziejski, D. McAdams, M. J. Podolsky, D. E. Fernandez, D. Farkas, and J. F. Christian, “A scalable correlator for multichannel diffuse correlation spectroscopy,” Proc SPIE Int Soc Opt Eng 9698, 969816 (2016).

41. J. X. M.D., X. H. M.D., Z. Qin, and F. Gao, “A multi-channel diffuse correlation spectroscopy system for dynamic topography of blood flow index in deep tissues,” 10.1117/12.2541810 11234, 115–125 (2020).

42. L. Kreiss, M. Wu, M. Wayne, S. Xu, P. McKee, D. Dwamena, K. Kim, K. C. Lee, K. R. Cowdrick, W. Liu, A. Ülkü, M. Harfouche, X. Yang, C. Cook, S. A. Lee, E. Buckley, C. Bruschini, E. Charbon, S. Huettel, and R. Horstmeyer, “Beneath the surface: revealing deep-tissue blood flow in human subjects with massively parallelized diffuse correlation spectroscopy,” Neurophotonics 12, 025007 (2025).

43. M. A. Wayne, E. J. Sie, A. C. Ulku, P. Mos, A. Ardelean, F. Marsili, C. Bruschini, and E. Charbon, “Massively parallel, real-time multispeckle diffuse correlation spectroscopy using a 500 × 500 SPAD camera,” Biomed Opt Express 14, (2023).

44. M. Wayne, A. Ulku, A. Ardelean, P. Mos, C. Bruschini, and E. Charbon, “A 500 × 500 Dual-Gate SPAD Imager with 100% Temporal Aperture and 1 ns Minimum Gate Length for FLIM and Phasor Imaging Applications,” IEEE Trans Electron Devices 69, (2022).

45. A. Gorman, N. Finlayson, A. T. Erdogan, L. Fisher, Y. Wang, F. M. Della Rocca, F. M. Della Rocca, H. Mai, H. Mai, E. J. Sie, F. Marsili, and R. K. Henderson, “ATLAS: a large array, on-chip compute SPAD camera for multispeckle diffuse correlation spectroscopy,” Biomedical Optics Express, Vol. 15, Issue 11, pp. 6499–6515 15, 6499–6515 (2024).

46. E. J. Sie, H. Chen, E.-F. Saung, R. Catoen, T. Tiecke, M. A. Chevillet, and F. Marsili, “High-sensitivity multispeckle diffuse correlation spectroscopy,” Neurophotonics 7, (2020).

47. W. Liu, R. Qian, S. Xu, P. Chandra Konda, J. Jönsson, M. Harfouche, D. Borycki, C. Cooke, E. Berrocal, Q. Dai, H. Wang, and R. Horstmeyer, “Fast and sensitive diffuse correlation spectroscopy with highly parallelized single photon detection,” APL Photonics 6, (2021).

48. Q. Wang, Y. Hua, C. Li, M. Pan, M. Wojtkiewicz, A. T. Erdogan, A. Gorman, Y. Zhang, N. Finlayson, Y. Wang, R. K. Henderson, and D. U.-D. Li, “Deep non-invasive cerebral blood flow sensing using diffuse correlation spectroscopy and ATLAS,” (2025).

49. K. R. Cowdrick, T. Urner, E. Sathialingam, Z. Fang, A. Quadri, K. Turrentine, S. Yup Lee, and E. M. Buckley, “Agreement in cerebrovascular reactivity assessed with diffuse correlation spectroscopy across experimental paradigms improves with short separation regression,” Neurophotonics 10, 025002 (2023).

50. Y. Gao, D. Rogers, A. von Lühmann, A. Ortega-Martinez, D. A. Boas, and M. A. Yücel, “Short-separation regression incorporated diffuse optical tomography image reconstruction modeling for high-density functional near-infrared spectroscopy,” Neurophotonics 10, 025007 (2023).

51. D. D. Duncan, S. J. Kirkpatrick, and E. M. Wells-Gray, “Detrimental effects of speckle-pixel size matching in laser speckle contrast imaging,” Optics Letters, Vol. 33, Issue 24, pp. 2886–2888 33, 2886–2888 (2008).

52. L. Cortese, G. Lo Presti, M. Pagliazzi, D. Contini, A. D. Mora, A. Pifferi, S. K. V. Sekar, L. Spinelli, P. Taroni, M. Zanoletti, U. M. Weigel, S. de Fraguier, A. Nguyen-Dihn, B. Rosinski, and T. Durduran, “Liquid phantoms for near-infrared and diffuse correlation spectroscopies with tunable optical and dynamic properties,” Biomed Opt Express 9, 2068 (2018).

53. D. Wang, A. B. Parthasarathy, W. B. Baker, K. Gannon, V. Kavuri, T. Ko, S. Schenkel, Z. Li, Z. Li, M. T. Mullen, J. A. Detre, and A. G. Yodh, “Fast blood flow monitoring in deep tissues with real-time software correlators,” Biomed Opt Express 7, 776 (2016).

54. C. H. Moore, U. Sunar, and W. Lin, “A Device-on-Chip Solution for Real-Time Diffuse Correlation Spectroscopy Using FPGA,” Biosensors (Basel) 14, (2024).

55. A. Biswas, P. P. S. Mohammad, S. Moka, A. Takshi, and A. B. Parthasarathy, “Non-invasive low-cost deep tissue blood flow measurement with integrated Diffuse Speckle Contrast Spectroscopy,” Frontiers in Neuroergonomics 4, 1288922 (2023).

56. Z. Li, W. B. Baker, A. B. Parthasarathy, T. S. Ko, D. Wang, S. Schenkel, T. Durduran, G. Li, and A. G. Yodh, “Calibration of diffuse correlation spectroscopy blood flow index with venous-occlusion diffuse optical spectroscopy in skeletal muscle,” J Biomed Opt 20, 125005 (2015).

57. S. Wojtkiewicz, M. Kacprzak, N. Mogharari, D. Borycki, and A. Liebert, “Time-domain diffuse correlation spectroscopy at large source detector separation for cerebral blood flow recovery,” Biomedical Optics Express, Vol. 15, Issue 7, pp. 4330–4344 15, 4330–4344 (2024).

58. R. C. Mesquita, S. S. Schenkel, D. L. Minkoff, X. Lu, C. G. Favilla, P. M. Vora, D. R. Busch, M. Chandra, J. H. Greenberg, J. A. Detre, and A. G. Yodh, “Influence of probe pressure on the diffuse correlation spectroscopy blood flow signal: extra-cerebral contributions,” Biomed Opt Express 4, 978 (2013).

59. N. Ozana, A. I. Zavriyev, D. Mazumder, M. Robinson, K. Kaya, M. Blackwell, S. A. Carp, and M. A. Franceschini, “Superconducting nanowire single-photon sensing of cerebral blood flow,” Neurophotonics 8, (2021).

60. J. Shao, “Consistency of least-squares estimator and its jackknife variance estimator in nonlinear models,” Canadian Journal of Statistics 20, 415–428 (1992).

61. J. Tellinghuisen, “Least Squares Methods for Treating Problems with Uncertainty in x and y,” Anal Chem 92, 10863–10871 (2020).

62. M. M. Wu, L. Kreiss, M. A. Wayne, M. B. Robinson, C. Bruschini, E. Charbon, and R. Horstmeyer, “Autocorrelation Bias in Diffuse Correlation Spectroscopy Observable via SPAD Arrays,” IEEE Journal of Selected Topics in Quantum Electronics 1–13 (2025).

